# Mapping and sequencing of cases from an ongoing outbreak of Clade Ib monkeypox virus in South Kivu, Eastern Democratic Republic of the Congo between September 2023 to June 2024

**DOI:** 10.1101/2024.09.18.24313835

**Authors:** Leandre Murhula Masirika, Jean Claude Udahemuka, Leonard Schuele, David F. Nieuwenhuijse, Pacifique Ndishimye, Marjan Boter, Justin Bengehya Mbiribindi, Cris Kacita, Trudie Lang, Christian Gortázar, Jean Pierre Musabyimana, Saria Otani, Frank M. Aarestrup, Freddy Belesi Siangoli, Bas B. Oude Munnink, Marion Koopmans, the GREATLIFE MPOX consortium

## Abstract

**Background:** In September 2023, an mpox outbreak was reported in the eastern part, South Kivu Province, of Democratic Republic of the Congo. This outbreak is still ongoing and expanding to other regions and countries. Here, we describe the epidemiological and genomic evolution of the outbreak from September 2023 to June 2024.

**Methods:** Consenting patients with mpox-like symptoms admitted to the Kamituga and the Kamanyola hospitals were recruited to the study. Samples from throat, lesions, breast milk and placenta were collected for PCR testing and sequencing. For the patients from Kamituga hospital, data on place of residence and possible exposures were collected by interviews. The location and numbers of employees were collected for all bars with sex workers. Where possible, exposures were linked to the genomic sequencing data for cluster analysis.

**Findings:** In total, 670 (suspected) mpox cases were admitted to the Kamituga hospital. There were slightly more female than male cases (351/670 [52,4%] versus 319/670, [47,6%], and cases were reported from 17 different health areas. The majority of cases were reported in Mero (205/670 [30,6%]), followed by Kimbangu (115/670 [17,2%]), Kabukungu (105/670 [16,7%]), and Asuku (73/670 [10.9%]). During this period, 7 deaths occurred and 8 out of 14 women who were pregnant had fetal loss. Three healthcare workers acquired mpox infection when caring for patients. In depth case ascertainment showed that 83,4% of patients reported recent visits to bars for (professional) sexual interactions as a likely source of infection. Whole genome sequencing resulted in the generation of 58 genome sequences. Three main clusters characterized by specific mutations were identified and several miniclusters of 2 or more sequences with over two shared mutations. No clear link between sequence cluster, bar or health area was observed. The more recent sequences from Kamanyola were related to the sequences in Kamituga and confirmed to be Clade Ib. However, relatively long branches were observed and one of the sequences clustered with publicly released sequences from travelers in Kenya, Uganda, Sweden and Thailand, indicating more undocumented ongoing spread for cluster A than for the other clusters. Most observed mutations were APOBEC-3 related mutations indicative of ongoing human-to-human transmission.

**Interpretation:** These data suggests that the rapid transmission of monkeypox virus until June 2024 was mostly related to interactions with professional sex workers (PSW) within densely populated health areas. The expanding number of cases and the recent expansion to 29 other nearby health zones of South -Kivu as well as Rwanda, Burundi, Uganda and Kenya stresses the need for cross border surveillance and collaboration. Urgent enhanced response action is needed, including case finding, diagnostic capacity building, health education programmes focussing on sex workers, and possibly vaccination to limit further escalation and stop this outbreak.

## INTRODUCTION

Human mpox disease (formerly known as monkeypox) is an emerging zoonotic disease caused by the monkeypox virus (MPXV). MPXV is a double-stranded DNA virus that belongs to the genus *Orthopoxvirus* within the *Poxviridae* family, subfamily *Chordopoxvirinae*. The first recorded human case of MPXV was in 1970 when a nine-month-old child was admitted to Basankusu Hospital in the Democratic Republic of the Congo (DRC) (Ladnyi et al,.1972). The clinical manifestations associated with MPXV resemble those of smallpox, a disease which has been globally eradicated since 1980. Infection in humans manifests as fever, swollen lymph nodes, and fatigue followed by a rash (genital, anorectal, or oral), with macular lesions progressing to papules, vesicles, pustules, and scabs, usually on the face, hands, and feet for two to five weeks Kaler et al.,(2022) ; MPXV is historically enzootic in Western and Central Africa and there have been sporadic mpox outbreaks in humans with increasing frequency primarily in West and Central African countries (Kaler et al., 2022). Though the exact host reservoir for MPXV is still unknown, some studies have shown that the reservoir is likely to be one or more species of squirrels or other rodents that inhabit the secondary forest of Central Africa (Curendeau et al., 2023; Doty et al., 2017; Kulesh et al., 2004). Since 2003, several cases of mpox have been reported in various countries. A study conducted between November 2005 and 2007 in DRC suggested that the incidence of human infections had massively increased since the cessation of smallpox vaccination programs, in line with a growing immunologically naive proportion of the population. The study also mapped increased risk to forested areas with an enzootic reservoir Rimoin et al., (2010).

The DRC is the sole country to have been continually reporting mpox cases in the last five decades, but there was limited global interest in supporting containment efforts. This changed when a widespread outbreak of mpox occurred worldwide in May 2022, with the first case confirmed in the United Kingdom in a man traveling from Nigeria Vaughan et al., (2022). This ongoing outbreak, caused by a clade II MPXV, is primarily transmitted through networks of men engaging in frequent unprotected sexual activities with other men (MSM) (Low et al., 2023; Wick et al., 2024). Although previous mpox outbreaks in the regions with enzootic MPXV reservoirs have suggested potential for spread to more diverse populations and through different modes of transmission, this has been very limited in the global outbreak.

Two genetically distinct clades of MPXV have been described: Clade I (formerly known as the Congo Basin or Central African clade) and Clade II (formerly known as the West African clade). Since August 2022, Clade II has been subdivided into two subclades, Clade IIa and Clade IIb. Viruses from Clade I and Clade II have a nucleotide sequence similarity of 99.4% Americo et al, 2023. MPXV Clade I is thought to cause more severe disease with reported case fatality rates (CFR) of up to 10.6%, while MPXV Clade II is associated with milder symptoms and a lower CFR of roughly 0.1%, but this is difficult to show with certainty given the importance of surveillance coverage and underlying health status on the disease course. https://www.who.int/news-room/questions-and-answers/item/monkeypox. Clade I is typically associated with (limited) transmission within households and often identifiable to a source of infection from bushmeat, with previously no or limited associations with sexual transmission. Since 2022, Clade II infections are primarily sexually transmitted. In November 2023 WHO noted an unprecedented increase in the number of notified cases across DRC, occurring across the region and including emergence in provinces that previously had not notified any cases https://www.who.int/emergencies/disease-outbreak-news/item/2024-DON522. Exact numbers are difficult to obtain, but in 2024, as of 15th of August, more than 16,000 cases have been notified in DRC with an estimated CFR of 3.4%. From limited genomic sequencing, it has become apparent that there are several ongoing outbreaks with different origins, age groups involved and suspected modes of transmission (Kinganda-Lusamaki et al., 2024). A particular concern has been the rapid increase of cases in South Kivu, since the initial detection in September 2023 with a shift in the epidemiological pattern compared to other regions (Katoto et al., 2024). Recently, we showed that the outbreak in South Kivu is associated with a novel sub-lineage of clade I viruses Murhula Masirika et al., (2024) and that the majority of these cases seem to be transmitted through (hetero)sexual contact.

Here, we provide a detailed overview of the outbreak in South Kivu from September 29th 2023 to the end of May 2024, including mapping of cases, assessment of possible exposures and virus genomic sequencing to understand the evolution of the outbreak.

## METHODS

### Study design and participants

We collected demographic data, and exposure histories of all individuals with MPXV infection symptoms admitted at the Kamituga Hospital between September 2023 and June 2024 who agreed to participate in the study. For consenting patients, geolocations were collected from the notification database in the health zones of South-Kivu province (https://data.humdata.org/dataset/drc-health-data). Ethical clearance to conduct these studies was obtained from the Ethical Review Committee of the Catholic University of Bukavu (Number UCB/CIES/NC/022/2023). All study participants were introduced to the observational study and given the option to participate by providing informed consent or, through parental permission. Patient information was anonymized and we confirm the IDs we used make the study participants unidentifiable.

### Study area

Kamituga is South Kivu’s largest gold mining city located in the territory of Mwenga, part of the South Kivu province. The South Kivu province borders in the north with the North Kivu province, south and west with the Maniema province, and south with the Tanganyika province. To the east, South Kivu borders Rwanda, Burundi, and Tanzania. The city of Kamituga has more than 241,642 inhabitants (based on the 2024 Kamituga Health Zone report), of which ∼20,000 are employed in the mining industry. The remaining residents depend directly or indirectly on artisanal and small-scale gold mining (ASGM) for their livelihood.

### Procedures and sample collection

Routine data like age, gender, professional occupation, clinical symptoms, geolocations of mpox case and concomitant presence of sexually transmitted infections (STIs), and comorbidities were collected from patient records (or hospital investigation forms) and entered into a secured, anonymized database. The study team was informed about the objectives of the study, the consent process, and the importance of standardized reporting.

Admission to the Kamituga Hospital was based upon clinical diagnosis of human MPXV infection by hospital staff. A confirmed MPXV case was defined as an individual with laboratory-confirmed infection which was tested in the National Institute for Biomedical for Research (INRB). A case was listed as “suspected” if a patient had an acute illness with fever, intense headache, myalgia, and back pain, followed by one to three days of a progressively developing rash often starting on the face and spreading on the body. Finally, a case was listed as “probable” if it satisfied the clinical definitions of suspected cases and had an epidemiological link to a confirmed or probable case but was not laboratory-confirmed. A skin lesion was defined as a single circumscribed area and included presentations comparable to papules, pustules, fluid-filled vesicles, or eschars.

### Mapping of mpox cases

Geographic and epidemiological data were processed using the R programming language. The epidemiological curve was plotted using the “ggplot2” package (Pebesma, 2018). For the geographical maps cases and sexworkers were scaled by population density and aggregated by health area defined by the shapefiles provided by national health ministry in Democratic Republic of the Congo available online: https://data.humdata.org/dataset/drc-health-data. Map plots were made by using the “sf” and “ggmap” R packages (Pebesma, 2018; Kahle and Wickham, 2013).

### Whole genome sequencing

A total of 92 samples from 88 patients covering the entire time-span of the ongoing mpox outbreak in the South Kivu region between September 2023 and May 2024 were selected for genomic investigations. Nucleic acids were extracted using the High Pure Viral Nucleic Acid Kit (Roche). A novel Clade Ib specific real-time PCR (Schuele et al., 2024), in combination with the US CDC generic MPXV real-time PCR (Li et al., 2010) were performed. Sixty-three samples below Ct 31 were selected for whole-genome sequencing (WGS). Two different amplicon-based approaches were utilized to ensure a high sequencing depth and coverage (https://github.com/pha4ge/primer-schemes mpxv/rigshospitalet/2500/v1.0.0; Brinkmann et al., 2022). Lysis buffer was used as negative control. Amplicon-based sequencing libraries were generated as described before (Murhula Masirika et al., 2024).

### Data analysis and generation of genomes

Raw sequence data of the two different amplicon-based approaches were preprocessed separately and merged afterwards to generate the final whole genome consensus sequences. Preprocessing was done by fastp with parameters “--disable_trim_polyg --disable_adapter_trimming --qualified_quality_phred 10 --unqualified_percent_limit 50 --length_required 1000” for the long (mpxv/rigshospitalet/2500/v1.0.0) or “--length_required 150” for the short (Brinkmann et al., 2022) amplicon panel given the different expected length of the amplicons. Initial primer trimming was performed by removing 30 nucleotides from both ends of the amplicon using cutadapt with parameters “-u 30 -u -30”. Additional primer trimming was performed using ampliclip (https://github.com/dnieuw/Ampliclip) for the long amplicons or by trimming an additional 30 nt at the beginning and end of the read for the short amplicon data since for the short amplicon data the exact primer positions are unknown. Reads were mapped to the reference genome NC_063383.1 with the parameters “-Y -x map-ont”. Coverage depth was calculated using samtools mpileup with parameters “-a -A -Q 0 -d 8000”. Variant calling was performed by generating a VCF file using our tool bam2vcf and parameters “-af 0.1” and filtering the VCF file to retain only major variants. Major variants were incorporated in the genome using a custom vcf2 consensus script and regions with coverage below the coverage threshold of 30x were masked with an “N”. A more detailed description of the analysis workflow and links to the tools is available at (https://github.com/dnieuw/mpox-south-kivu-mapping-manuscript). Additional manual curation was done by verifying mutations that were only present in either the long or the short amplicon set data and by masking for mutations in homopolymer or di, tri or multimer repeat regions. Sequences with >85% genome coverage (n=58) were included in phylogenetic analysis.

All available clade I sequences on NCBI and GISAID on the 31st of August 2024 were merged and a maximum likelihood phylogenetic tree was generated using IQ-Tree using the K3Pu+F+I model as best predicted model (GISAID acknowledgment is present Supplement Table 1). The maximum likelihood phylogenetic tree was visualized using a custom R script making use of mainly the ggtree, tidytree, ape, and patchwork packages. APOBEC3 mutations were identified as described by O’Toole et al., 2023. Based on the tree and the specific mutations, we identified clusters with arbitrarily assigned numbers that were subsequently added as a field to the metadata file for testing of possible associations with potential exposure routes (specific bars, households, etc).

**Table 1:** Description of number of inhabitants, bars, persons employed as sexworkers, notified mpox cases for the health areas that notified cases during the study period. Last column indicates from which area samples were successfully sequenced.

| Health Zone | Health areas | Case | % | PSW | Bars | Population | Sequenced |
| --- | --- | --- | --- | --- | --- | --- | --- |
| Kamituga | KELE SIDEM | 10 | 1,49 | 0 | 0 | 10618 |  |
| Kamituga | ASUKU | 73 | 10,90 | 38 | 3 | 11002 | 8 |
| Kamituga | BIGOMBE | 3 | 0,45 | 18 | 2 | 7274 |  |
| Kamituga | ISOPO | 1 | 0,15 | 0 | 0 | 5403 |  |
| Kamituga | KABUKUNGU | 115 | 17,16 | 152 | 10 | 13758 | 7 |
| Kamituga | KALINGI | 28 | 4,18 | 57 | 5 | 12888 | 4 |
| Kamituga | KANKANGA | 6 | 0,90 | 0 | 0 | 5622 | 1 |
| Kamituga | KATUNGA | 43 | 6,42 | 7 | 1 | 13492 | 3 |
| Kamituga | KELE KATUTU | 3 | 0,45 | 21 | 0 | 11234 |  |
| Kamituga | KIBE | 2 | 0,30 | 34 | 3 | 12073 |  |
| Kamituga | KIMBANGU | 105 | 15,67 | 45 | 5 | 16132 | 11 |
| Kamituga | LULIBA | 11 | 1,64 | 18 | 2 | 2870 | 1 |
| Kamituga | MERO | 205 | 30,60 | 161 | 13 | 25598 | 14 |
| Kamituga | NGAMBWA | 2 | 0,30 | 15 | 0 | 4025 |  |
| Kamituga | POLY AFIA | 13 | 1,94 | 78 | 5 | 12105 | 3 |
| Kamituga | POUDRIERE | 14 | 2,09 | 32 | 6 | 12931 |  |
| Kamituga | SOLULUYU | 36 | 5,37 | 75 | 8 | 10921 | 4 |
|  | **Total** | **670** | **100,00** | **751** | **63** | **187946** |  |
| Nyangezi | KAMANYOLA |  |  |  |  |  | 2 |

## RESULTS

### Trends in hospitalized cases within health areas of Kamituga

A total of 670 hospitalized mpox case records were listed as confirmed, probable, or suspected **(Figure 1A)** between September 29th, 2023 and June 30th, 2024. Three of these were health care workers, and seven deaths occurred during the study period (1%). In contrast to other outbreaks in the DRC, only 15,5 % (104/670) of suspected cases were children under 15 years of age. Of these, 45 were less than 5 years old. The majority of patients were aged between 16 -26 years old. In total, 646/670 (96.1%) were confirmed by PCR. There were slightly more female than male cases (351/670 [52,4%] versus 319/670, [47,6%], and cases were reported from 17 of the 23 health areas (Table I). In total, 7 persons died during the study period (1%). The majority of cases were reported in Mero (205/670 [30,6%]), Kimbangu (115/670[17,2%]), Kabukungu (105/670[15,7%]), and Asuku (73/670 [10.9%]) (Table I).

**Figure 1.**
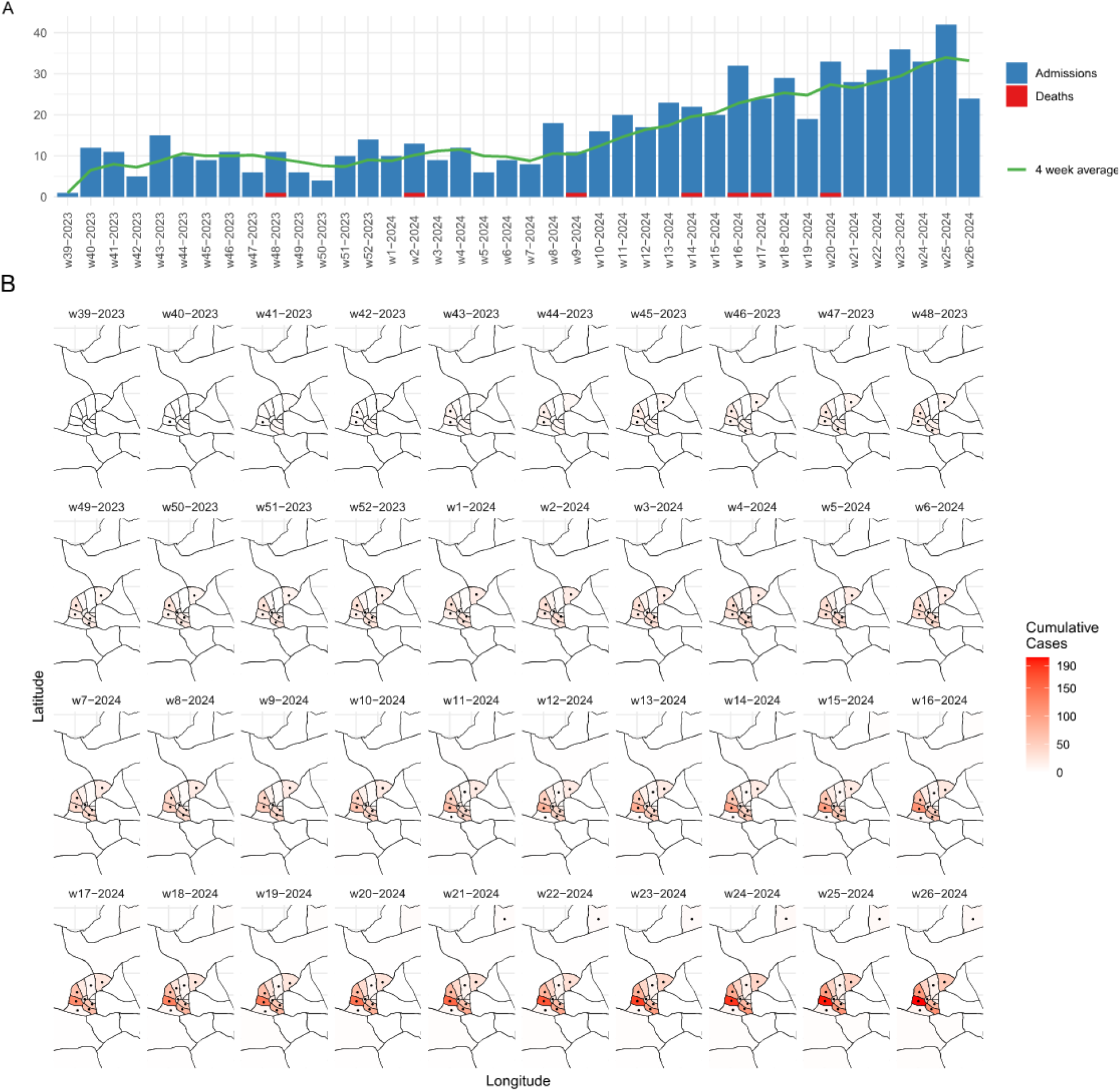
**(A)** Epicurve of outbreak based on Kamituga health zone surveillance dataset for the mpox patients’ admissions at Kamituga hospital and the deaths per epidemiological week between September 29th, 2023 to June 29th, 2024. **(B)** Map showing evolution of the outbreak in time and space within health areas of Kamituga health zone. Black dots indicate in what epidemiological week and health area a first complete genome sequence was generated.

The highest number of admissions in a single week for the year 2023 was 14, reported in weeks 43, 48, and 52 in the Kamituga health zone. In 2024, the number of mpox cases sharply increased with the highest number of 42 admissions reported in week 25. The 4-week moving average also increased from around 10 hospitalized cases per week in 2023 with a gradual increase to over 30 cases hospitalized per week in the Kamituga hospital since the beginning of 2024 **(Figure 1A, Figure 1B)**. The outbreak also expanded to neighboring health areas, namely Bigombe, Luliba, Isopo, Kele Katutu, Ngambwa and Kankanga, respectively, in the epidemiological weeks 7, 10, 11, 15 and 24 of the year 2024. In epidemiological week 41 of 2023, a few cases were identified in Nyangezi health zone, which is bordering Rwanda and Burundi. From week 14 in 2024 onwards, the number of notifications from this region steeply increased. Given this expansion, a few cases were sampled to include in the sequencing analysis of the Kamituga hospital patient selection.

### Description of fatal cases

Four of the seven fatalities were young adults (between 20 and 30 years of age), with 3 females and 1 male patient. One patient had HIV co-infection and died after developing neurological complications, and a second patient had complications from a cesarean section. A third person developed confluent lesions, with clinically suspected bacterial infection, and patient number four died after developing respiratory distress. The other three recorded deaths were infants, one that was born with lesions from intra-uterine infection, and two babies infected post-natally. In two cases breast milk samples tested positive. HIV status was not known. As there was no clinical characterisation protocol in place, further details are not available, but since March 2024, a cohort study was initiated for more systematic case ascertainment. This work is currently ongoing.

In addition to these, severe disease outcomes were observed in pregnant women. During the study period, 14 pregnant women were hospitalised, of whom eight (57,1%) suffered intra-uterine fetal death and or fetal loss, with visual evidence of infection (pox-lesions) in 1 fetus and a PCR positive placenta in another case.

### Demographics of health areas and assessment of number of cases, bars and professional sex workers within bars per health area

On September 29th 2023, the index mpox case was reported in the Poudriere health area, and was linked to a bar visit. We interviewed hospitalized cases with a standardized case reporting form including assessing potential sexual exposures. Of the 670 hospitalized mpox cases, 83.4% (559/670) reported recent sexual contact in bars among which 44,6 % were female and 38.8% were male. Only a few cases reported to not have had sexual contact in bars: in total 16.6% (111/670) with 8.8% (59/670) males and 7.8 % (52/670) females. The majority of these were reported to be contacts of known cases, but there is insufficient information for a more detailed breakdown of possible exposures in this group of individuals. Following the initial observation of the contribution of sex workers driving transmission, and the mentioning of specific bars as possible places of exposure, we carried out a census of bars and professional sex workers (PSW) of Kamituga health zone (Table I). Kamituga mining city has 63 bars distributed as shown in Table I. We also inventoried the number of PSWs working in bars (751 in total) within Kamituga health zones. The highest numbers were reported in Mero health area with 21% (161/751) followed by Kabukungu with 20% (152/751), Poly Afia 10 % (78/751), Soluluyu 10% (75/751), Kalingi 8% (57/751), Kimbangu 6% (45/751), Asuku 5% (38/751), Kibe 5% (34/751), Poudriere 4% (32/751), Kele Katutu 3% (21/751), Bigombe 2% (18/751), Luliba 2 % (18/751), Mulambula 2% (15/751), and Katunga 1% (7/751) **(Figure 2 B)**.

**Figure 2:**
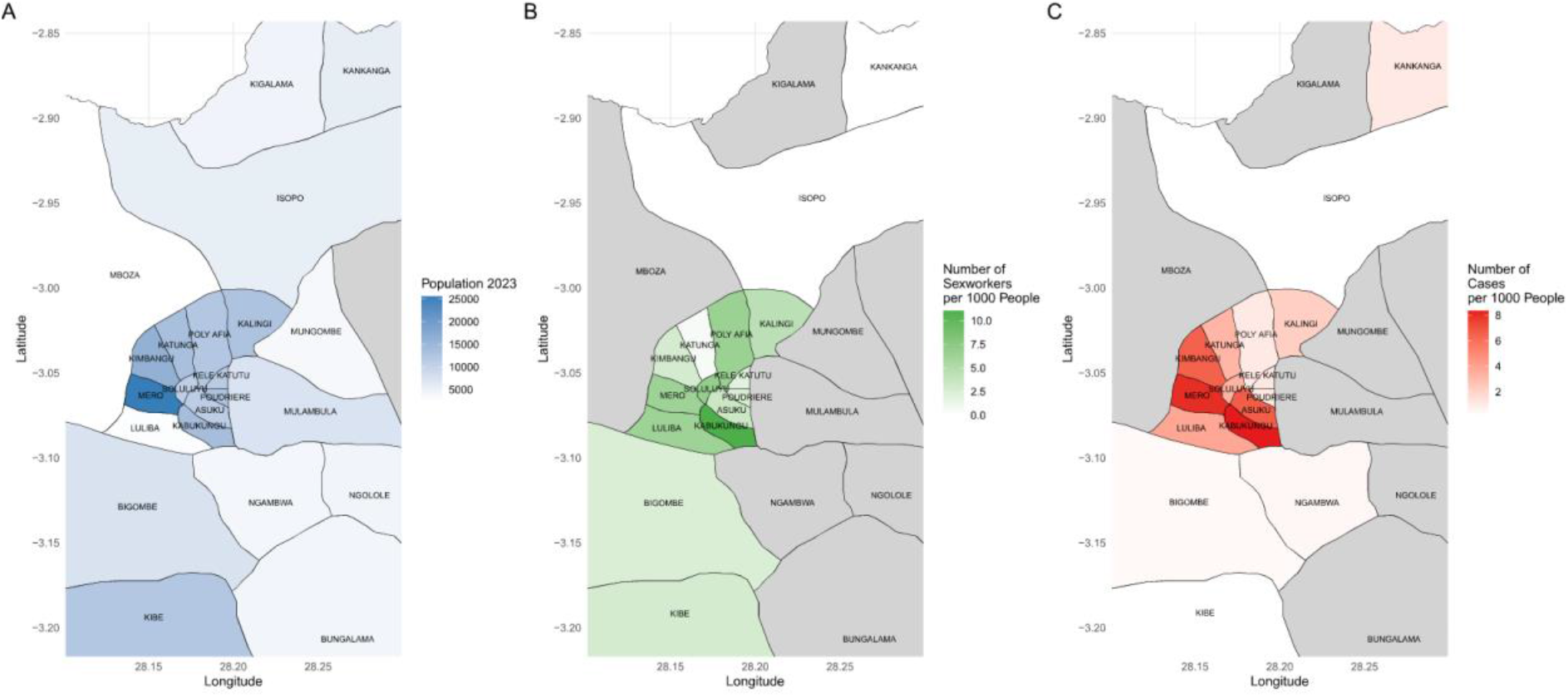
**(A)** reports population density (2023) per health area in Kamituga health zone, between September 29th, 2023 and April 21st, 2024, South-Kivu, DRC, **(B)** reports number of sex workers per 1000 inhabitants, **(C)** reports number of mpox cases per 1000 inhabitants. Gray areas do not have any recorded population (A), sex workers (B), or have not reported any cases (C).

**Figure 3.**
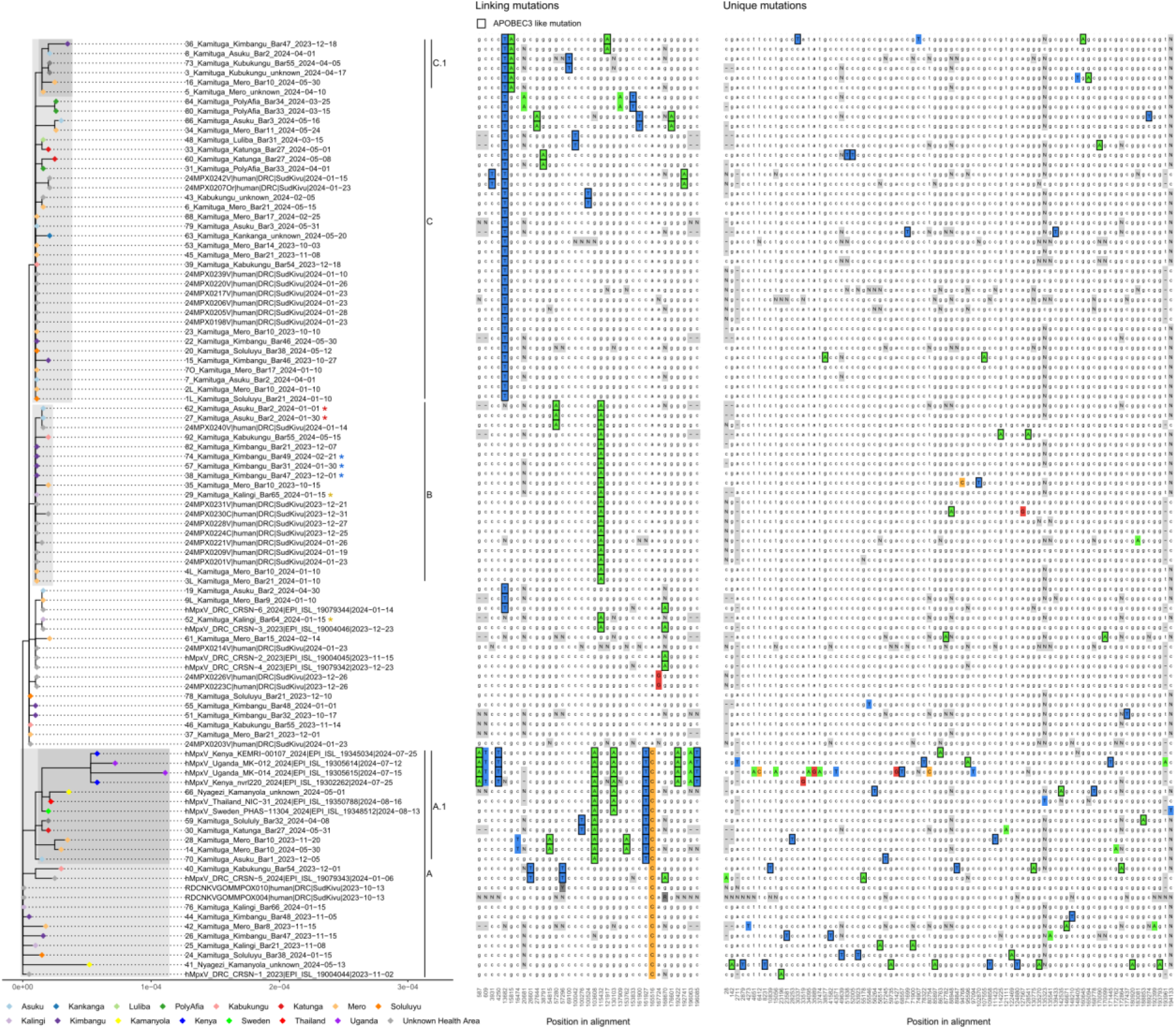
Zoom-in of phylogenetic tree specific for the currently shared and newly sequenced clade Ib sequences. Left panel shows the phylogenetic tree with colored tips indicating health area of originating patient. Colored asterisks indicate sequences from different samples of the same patient. Proposed clusters are shown by shading and annotated by markings on the right side of the tree. Middle panel shows linking mutations, present in two or more cases. Right panel shows unique mutations. Differences from the majority rule consensus are highlighted in color. Mutations with characteristic APOBEC3 signature are marked with a black box. Positions that were masked by manual curation or due to too little coverage are indicated with “N”. Ambiguous nucleotides are marked in dark gray.

We mapped the population density of the Kamituga health zone, as well as the number sex workers and reported mpox cases during the study period (Figure 2). In 2023 Mero had the highest population density, followed Kimbangu and then by six health areas with similar numbers of people (Kalingi, Poly Afia, Kibe, Poudriere, Katunga, and Kabukungu; Table 1, Figure 2A). As expected, the expansion of the outbreak from September onward closely matched these more populated health areas (**Figure 2C**).

### Phylogenetic analysis of the generated whole genome sequences

We sequenced MPXV genomes from samples during the current outbreak collected between October 2023 and May 2024. Fifty-eight near-complete whole genome sequences were generated from 54 patients. Phylogenetic analysis and inspection of the mutation patterns revealed three potential clusters and two potential subclusters consisting of sequences of different collection dates including April/May 2024, suggesting several different, ongoing, transmission chains. Indicated by the colored tips, the cluster patterns did not directly relate to the health area origin of the patients. Cluster A shows many unique mutations suggesting undetected circulation or circulation outside the Kamituga health zone, also corroborated by the two samples in this cluster from Kamanyola (yellow tips). Cluster B shows little shared and unique mutations compared to the other clusters. This cluster shows several cases of “miniclusters” of two cases with identical sequences. Three of these mini clusters consisted of sequences from the same patient (27 and 62, 29 and 52, and 38, 57 and 74 are sequences from the same patient).

We observed an abundance of APOBEC3 mutations (28/35 linking mutations and 55/82 unique mutations) indicative of human-to-human transmission. Most APOBEC3 mutations were observed in cluster A, which includes the two sequences from cases from Kamanyola. These two cases are not directly linked to each other and show up to 10 unique mutations, suggesting considerable transmission prior to their detection. The sequences of travel associated cases in Kenya, Tanzania, Thailand and Sweden are part of sub-cluster A.1 and show an additional 7 linked mutations, of which 5 have the APOBEC3 signature, however 6 of the 7 mutations are situated in the inverted terminal repeat region effectively creating a reverse complement duplication of the mutations.

## Discussion

Few formal case investigations of Clade I have been documented during outbreaks in Central Africa, and no significant human-to-human transmission of Clade I cases was reported until April 2023. The first documented case of the sexually transmitted MPXV Clade Ia virus involved a male resident of Belgium who had traveled in the Democratic Republic of the Congo and had sexual encounters in Kwango province (southwest) (Kibungu et al., 2024). Prior to 2023, South Kivu Province had not reported any continuous mpox cases, except for the limited sporadic mpox cases reported in Shabunda territory, South-Kivu in 2014 (McCollum et al., 2015). Recently, a new mpox sublineage was identified causing the current outbreak in South Kivu, Clade Ib, with suggestive evidence for a different route of transmission of the virus, primarily through (hetero)sexual contact (Murhula Masirika et al., 2024; Vakaniaki et al., 2024). The data we have compiled here provides additional insights into the time trend and spatial distribution of the ongoing mpox outbreak in South Kivu, DRC, and sheds light on the links of increasing mpox cases with population density and the presence of PSWs in bars within affected heath areas.

The health areas reporting continuous mpox cases were in densely populated areas with a high number of bars, that are sustaining the local sex industry. These activities are related to gold mining, which is a major activity in the region and has attracted an extensive sex worker industry. The majority of the patients reported to have had contact with bars in the two or three weeks before they started to develop mpox-like symptoms. Since the start of the outbreak, the second highest number of mpox cases was reported from the Nyangezi health zone, which has similar activity of mining including the city of Kamanyola which is rich in cassiterite mining, and is located at the crossroads of three countries, DRC, Rwanda and Burundi. People coming from remote mining areas spend the night in Kamanyola city and possibly in bars before crossing either to Rwanda, Burundi or to other nearby towns in DRC such as Bukavu and Goma. PSWs from Burundi, DRC and Rwanda work in bars in Kamanyola, pointing at an increased risk for cross-border transmission. Of note Rwanda, Uganda, Kenya, Sweden and Thailand have recently reported cases of mpox by Clade Ib variant, linked to people traveling from DRC, while Burundi also has reported sustained transmission within the country of clade Ib since the 25th of July 2024 (https://www.who.int/emergencies/disease-outbreak-news/item/2024-DON528).

There currently is considerable uncertainty about properties associated with the new Clade Ib variant, regarding transmissibility and severity of disease associated with infection. Based on animal studies and outbreak investigations, virulence differences have been observed between Clade I and Clade II virus infections, although this data is difficult to state with certainty due to the challenges in case ascertainment in low-income settings, and the lack of systematic follow-up of cases in households (Okwor et al., 2023). In this study, a case fatality rate of 1% was observed, which likely is an overestimate given these were hospitalised patients. Case fatality rate may also be influenced by the age range of cases; we observed 3 deaths among 45 children less than 5 years of age. In addition, the deletion of the virulence gene C3L in the clade Ib viruses, which is also not present in the clade II viruses, might result in lower virulence but further studies are needed to elucidate this.

A specific concern is the potential for complications of mpox in pregnant women. During this outbreak, 14 pregnant women were admitted at Kamituga hospital among whom 8 aborted, supporting the potential for intrauterine transmission of MPXV in pregnant people as also previously suggested (Cuérel et al., 2022; Schwartz et al., 2008). In 2008 a MPXV Clade I case of fetal death after placental infection was reported in at the Kole Hospital in Kole, located in the Sankuru District of Kasaï-Oriental Province, which findings suggested risks of intrauterine infection emphasizing that clinicians should be aware of this potential complication (Schwartz et al., 2008). We also show the detection of MPXV DNA in breast milk samples, showing the potential of mother-to-child transmission during breastfeeding. However, the current knowledge precludes a separate assessment of the role of milk versus direct contact in transmission of MPXV from mothers to infants.

A further knowledge gap is the question if there are differences in transmissibility between mpox clades, and between Clade Ia and Ib viruses. While there is suggestive evidence from the increased rate of person-to-person spread observed in Clade Ib infections, the opportunity for spread provided by the specific risk behavior may also explain the observed pattern. The proportion of patients reporting recent exposure to a risk environment (bars, prostitution) has not decreased over time, providing some evidence that the outbreak still is primarily driven by close sexual contact. A recent animal study found an association between routes of exposure, viral load, and rate of secondary transmission, with increased shedding and more transmission in animals infected by vaginal or rectal inoculation (Port et al, 2024).

The whole genome sequencing data was added to provide further insight into the Clade Ib dispersal. Several clusters and potential subclusters could be identified which can facilitate tracking the spread of the current outbreak. Several APOBEC3 signature mutations were observed, particularly in cluster A sequences. Based on currently available public data all cross-border cases to date are linked to this cluster, suggesting sustained but undetected human-to-human transmission of clade Ib monkeypox virus outside the Kamituga health zone before reaching Kamanyola.

Important unknowns remain and we are continuing our programme of studies. These seek to assess the extent of transmission among sexworkers and household members of known cases, for which serological testing will be included.

## Conclusion

In conclusion, we report an ongoing mpox outbreak expanded to over 17 Kamituga zone’s health areas, thus far, causing 7 deaths among 670 persons admitted to the Kamituga hospital. In addition, 8 out of the 14 pregnant women infected with mpox aborted. The outbreak appears to be driven by sexual activity with PSWs linked to bars supporting the previously undocumented model of heterosexual transmission. Genomic analysis shows evidence for considerable underdetection. Cross border surveillance, access to rapid diagnostics and treatment, and potential vaccination are crucial to control further dispersal.

## Supporting information

Figure 2 :(A) reports population density (2023) per health area in Kamituga health zone, between September 29th, 2023 and April 21st, 2024, South-Kivu

Figure 3: Zoom-in of phylogenetic tree specific for the currently shared and newly sequenced clade Ib sequences. Left panel shows the phylogenetic tre

## Author Approval

All authors approved the final version of the manuscript. LMM, JCU, PN, MK, conceptualized and designed the study. LMM, BBOM, LS, MB, MK, FMA, SO, DN, contributed to data acquisition, LMM, BBOM, LS, DN, MK drafted the manuscript and figures. LMM, JBM, FBS collected epidemiological data and arranged for patient sampling. All authors were involved in manuscript writing and approved the final version.

## Ethical approval

The ethical clearance to conduct these studies was obtained from the Ethical Review Committee of the Catholic University of Bukavu (Number UCB/CIES/NC/022/2023.

## Data availability

Consensus sequences are available on the GISAID under the accession numbers: xx-xx and on GenBank under the accession numbers: PQ305763-PQ305820. Raw sequence data is available on the ENA under the accession numbers: xx-xx.

## Acknowledgements

We would like to thank the Provincial Division of Health (DPS) of South-Kivu and Kamituga Health Zone (KHZ) for their collaboration during the study. We greatly thank the National Biomedical Research Institute (INRB) for mpox cases confirmation during the ongoing outbreak.

