## Supplementary figures and images for "Mapping and sequencing of cases from an ongoing outbreak of Clade Ib monkeypox virus in South Kivu, Eastern Democratic Republic of the Congo between September 2023 to June 2024"

### Figure 2 :(A) reports population density (2023) per health area in Kamituga health zone, between September 29th, 2023 and April 21st, 2024, South-Kivu

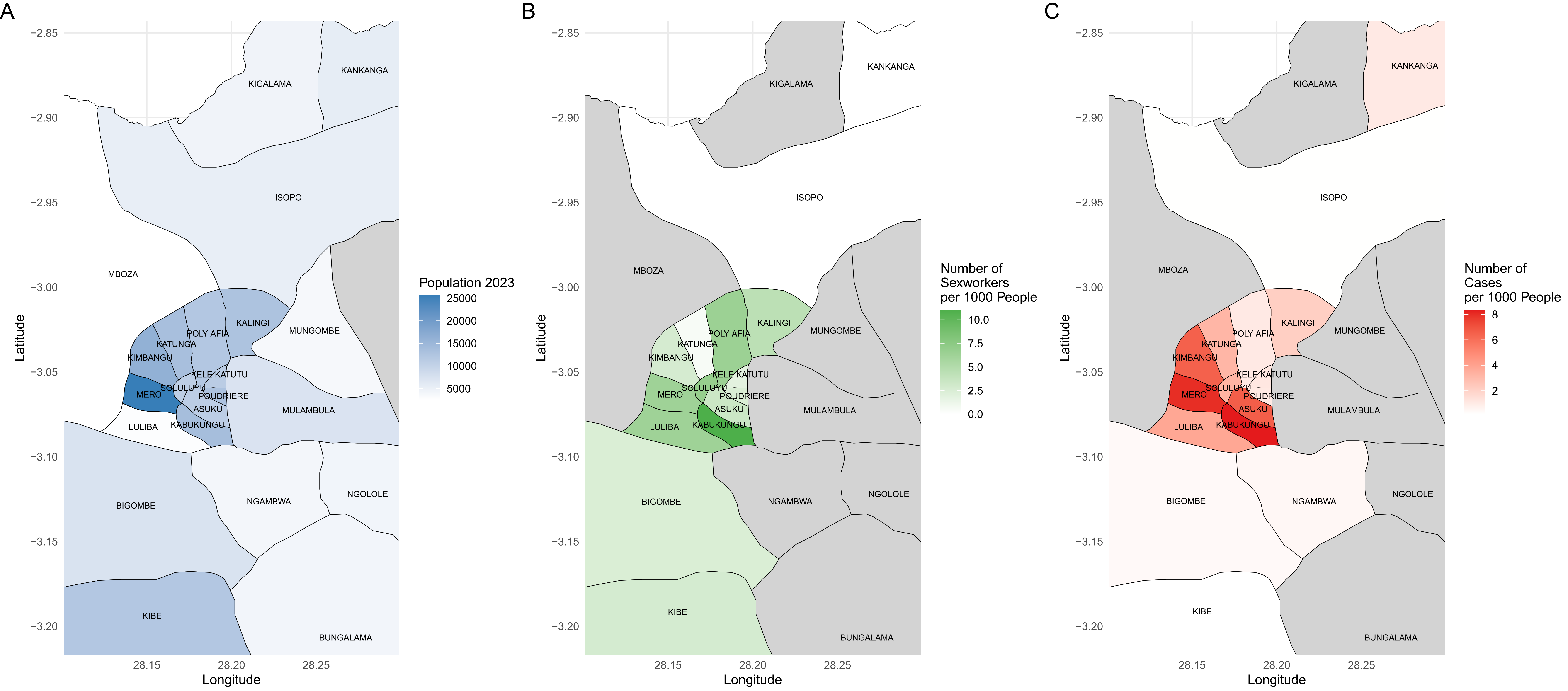

### Figure 3: Zoom-in of phylogenetic tree specific for the currently shared and newly sequenced clade Ib sequences. Left panel shows the phylogenetic tre

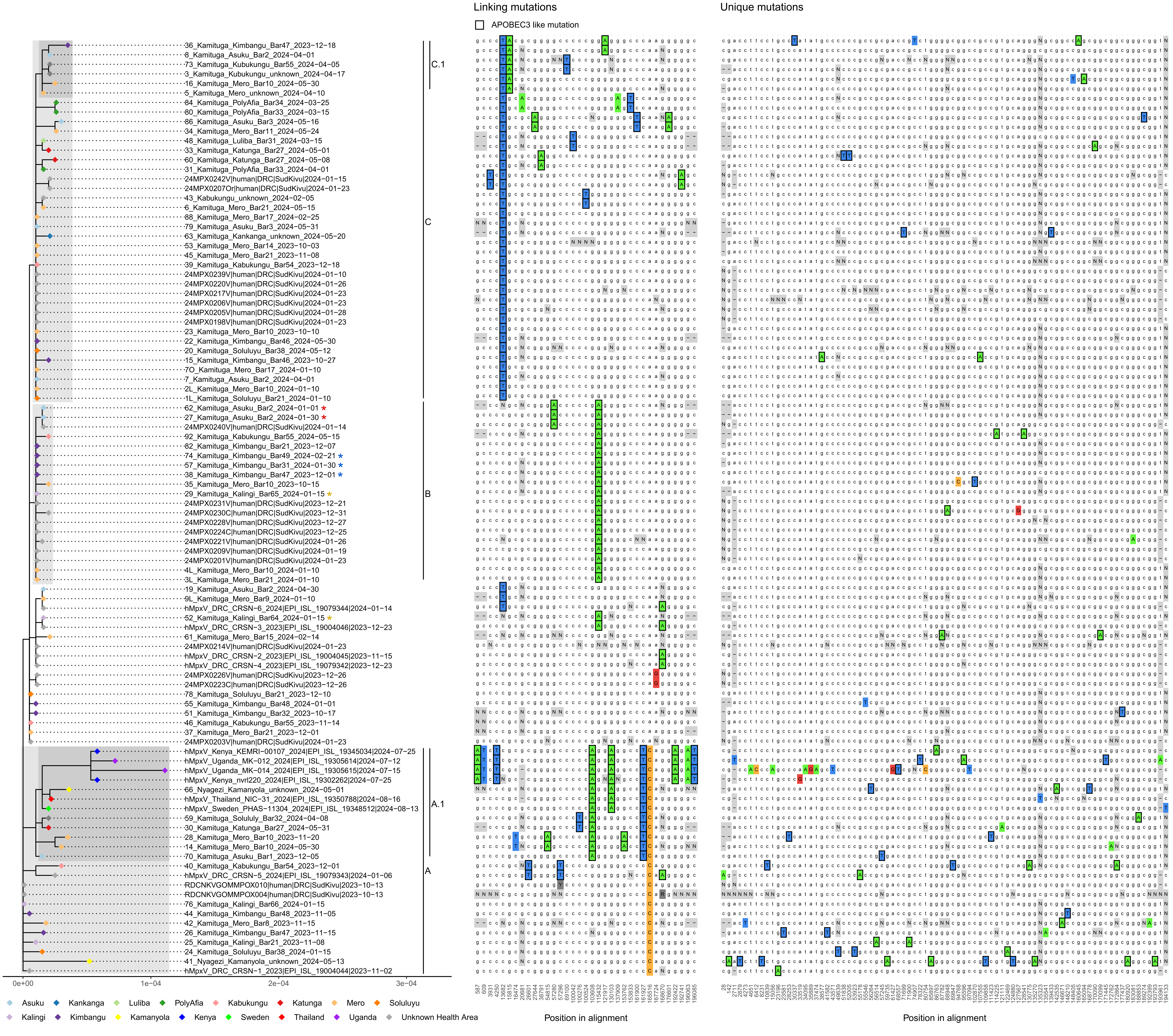

### Supplemental Data 1

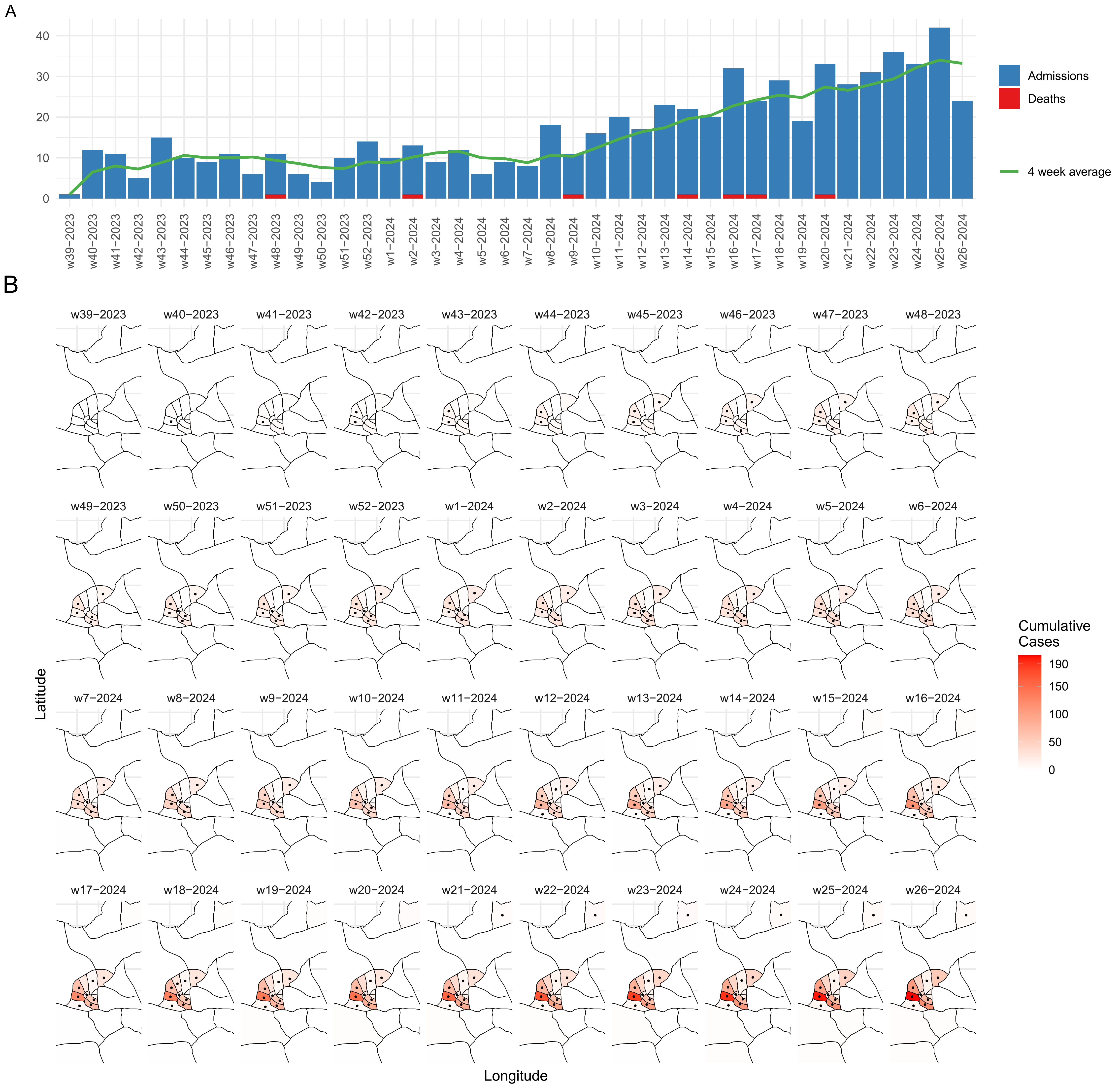
